# Verifiable Summarization of Electronic Health Records Using Large Language Models to Support Chart Review

**DOI:** 10.1101/2025.06.02.25328807

**Authors:** Ritchie Verma, Emily Alsentzer, Zachary Strasser, Leslie Chang, Kirollos Roman, Esteban Gershanik, Camellia Hernandez, Miguel Linares, Jorge Rodriguez, Durga Thakral, Ozan Unlu, Jacqueline You, Li Zhou, David Bates

## Abstract

Information overload in electronic health records (EHRs) hampers clinicians’ ability to efficiently extract and synthesize critical information from a patient’s longitudinal health record, leading to increased cognitive burden and delays in care. This study explores the potential of large language models (LLMs) to address this challenge by generating problem-based admission summaries for patients admitted with heart failure, a leading cause of hospitalization worldwide. We developed an extract-then-abstract approach guided by disease-specific “summary bundles” to generate summaries of longitudinal clinical notes that prioritize clinically relevant information. Through a mixed-methods evaluation using real-world clinical notes, we compared physicians’ ability to answer patient-specific clinical questions with the LLM-generated summaries versus standard chart review. While summary access did not significantly reduce overall questionnaire completion time, frequent summary use significantly contributed to faster questionnaire completion (p = 0.002). Individual physicians varied in how effectively they leveraged the summaries. Importantly, summary use maintained accuracy in answering clinical questions (88.0% with summaries vs. 86.4% without). All physicians indicated they were “likely” or “very likely” to use the summaries in clinical practice, and 87.5% reported that the summaries would save them time. Preferences for summary format varied, highlighting the need for customizable summaries aligned with individual clinician workflows. This study provides one of the first extrinsic evaluations of LLMs for longitudinal summarization, demonstrating their potential to enhance clinician efficiency, alleviate workload, and support informed decision-making in time-sensitive care environments.

## Introduction

A comprehensive understanding of a patient’s medical history at hospital admission and during their stay is vital for accurate diagnosis, informed clinical decision-making, and the development of personalized treatment plans. However, information overload in electronic health records (EHRs) makes it difficult for clinicians to identify salient clinical information from a patient’s longitudinal health record, which can contain hundreds to thousands of clinical notes. Chart review in the EHR requires clinicians to switch tasks frequently and transition between screens to gather patient information^1^. This time-consuming and tedious process of summarizing a patient’s medical history, especially during note generation, can result in errors in documentation and delays in diagnosis and treatment, impacting the quality and effectiveness of patient care^2–5^. Moreover, the substantial workload associated with documentation and other administrative tasks has been implicated in reduced career satisfaction and heightened rates of burnout among healthcare providers,^6^ which are at high levels.

Automatic summarization has significant potential to improve the efficiency and quality of chart review by reducing cognitive load and enabling clinicians to synthesize critical information quickly. Several tools have been developed to address this challenge by enabling visualization of patient histories^7–10^. For example, HARVEST provides a problem-based temporal visualization of longitudinal patient records, organizing clinical data on a timeline and linking problems to the corresponding clinical notes where they are mentioned^7^. MedKnowts integrates note-taking with information retrieval, offering concise syntheses of clinical concepts mentioned in the note, including relevant labs, imaging, and links to specific notes^10^. Other tools leverage natural language summaries of patient data^11–13^. For example, the BabyTalk project includes two tools to generate nursing shift summaries from structured data from the neonatal intensive care unit over a period of 45 minutes (BT-45) and 12 hours (BT-Nurse)^11^. While these tools provide valuable innovations, they do not fully address the complexity of synthesizing longitudinal patient records. Existing tools excel at organizing or retrieving information and generating summaries from structured datasets, but fall short of providing comprehensive summaries for longitudinal, heterogeneous, and unstructured patient records^14^.

Recent large language models (LLMs), such as GPT-4, have demonstrated immense promise in summarization tasks across the clinical domain. LLMs have been leveraged to generate discharge summaries^15–20^, summarize progress notes and radiology reports^21^, and summarize medical evidence^22^. Furthermore, clinical summarization is included in a major EHR vendor’s (Epic, Verona, Wisconsin) strategy for AI tool implementation^23,24^. However, the potential for longitudinal summarization of patient medical records to support clinicians performing chart reviews remains underexplored. Unlike previous applications focused on single hospital admissions or individual clinical notes, chart review upon hospital admission involves synthesizing a patient’s extensive longitudinal medical record. This task presents unique challenges for LLMs, which must be able to address inconsistencies and variations in the reliability of information across historical clinical notes to generate a coherent summary. Furthermore, the substantial length of longitudinal records for many patients exceeds the context window of current LLMs, necessitating strategies to prioritize relevant information efficiently.

In this paper, we evaluated the utility of LLMs for creating problem-based admission summaries using clinical notes from the EHR. We focused on patients with heart failure (HF), one of the most common diagnoses for inpatient hospitalization in the United States.^25^ To develop reliable and clinically grounded summaries, we leveraged domain expertise to create summary bundles^26^: standardized templates specifying the key elements of a patient’s disease history to include in each summary. These bundles serve as structured frameworks for extracting pertinent clinical information from the patient’s chart. Using the summary bundles, we implemented a two-step approach to build longitudinal, problem-based summaries. First, we leveraged LLMs to perform zero-shot extraction of information specified in the summary bundles from notes prior to admission. Then, we summarized the extracted information into a coherent summary of the patient’s prior medical history. Our domain expert–guided “extract-then-abstract” approach enables precise selection of pertinent clinical details while ensuring that each sentence in the summary is traceable to a source note, supporting more reliable and interpretable outputs. To evaluate this approach, we conducted an extrinsic assessment by comparing physicians’ ability to answer patient-specific clinical questions using LLM-generated summaries versus standard chart review processes. Unlike prior evaluations that focus solely on the intrinsic quality of summaries, our approach assessed how summaries impact physicians’ ability to perform chart reviews.

## Results

### Study Overview

When evaluating patients presenting for hospital admission, clinicians often lack prior familiarity with the patient and must quickly synthesize extensive longitudinal records to understand their medical history, identify relevant clinical issues, and develop a plan for admission. This pre-admission process is complicated by the volume and complexity of electronic health records (EHRs), which can include hundreds or thousands of clinical notes, making the task both time-consuming and cognitively demanding. To address this challenge, we developed problem-based longitudinal summaries using large language models (LLMs) to provide disease-oriented overviews tailored to facilitate the admission process. These summaries aim to streamline chart reviews by reducing the time required to identify and synthesize salient clinical information while maintaining relevance and accuracy.

Our patient population included individuals seen at Brigham and Women’s Hospital between 2017 and 2023, with a focus on patients admitted for heart failure. HF is a leading cause of hospitalization in the United States, accounting for over 1 million hospital admissions annually^27,28^. Its chronic nature and the need for detailed longitudinal assessments to guide treatment decisions make HF an ideal test case for evaluating the feasibility and utility of LLM-generated summaries. By focusing on HF, we sought to establish a framework for generating and evaluating problem-based summaries, providing a foundation for expanding this approach to other medical conditions.

### Overview of Verifiable Summarization Approach

We developed a structured, verifiable process to generate longitudinal summaries for heart failure patients, leveraging domain-specific knowledge and a two-step summarization framework to ensure accuracy, comprehensiveness, and traceability (Figure 1).

**Figure 1:**
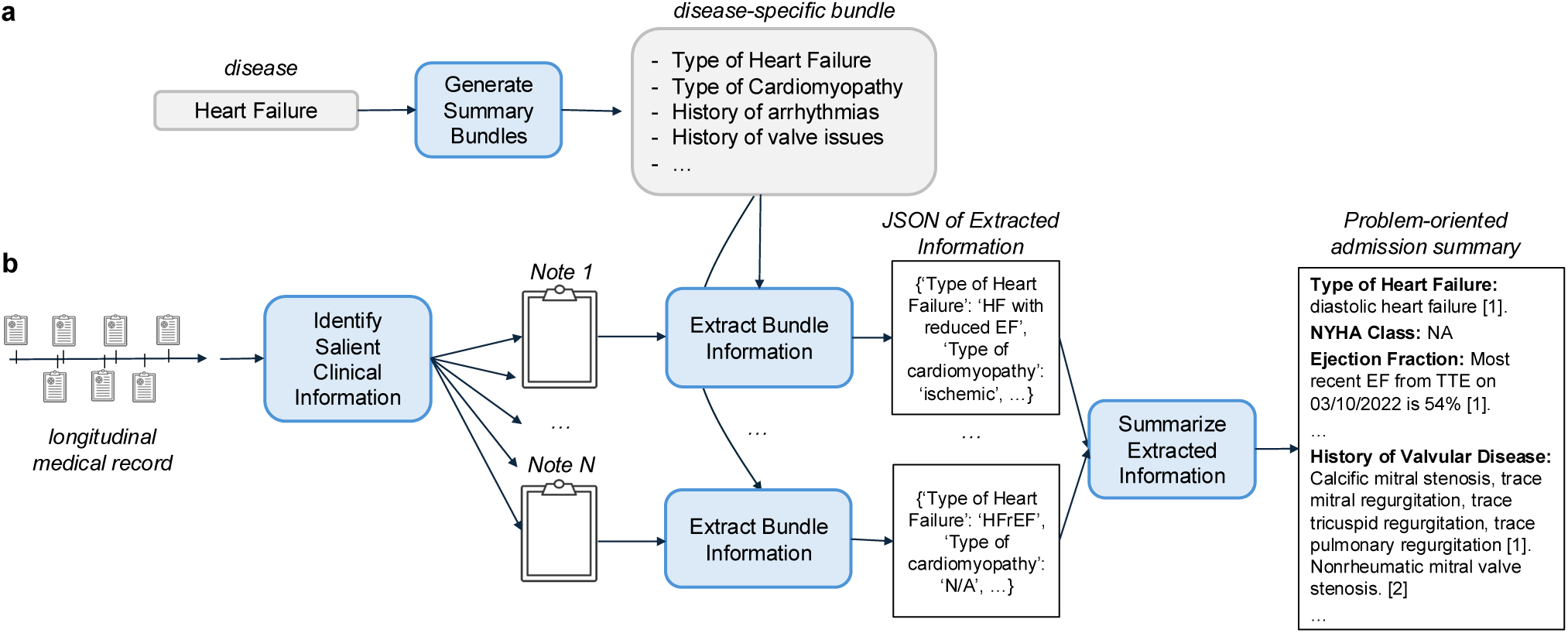
Overview of the summarization pipeline. (a) Domain expertise is leveraged to develop disease-specific bundles, which define key clinical data elements essential for managing hospitalized heart failure (HF) patients. (b) Summaries are generated using a two-step “extract-then-abstract” approach. In the first step, relevant information is extracted from individual patient notes based on the predefined bundles. In the second step, these extracted details are synthesized into a cohesive, structured summary.

Summaries were constructed using notes recorded prior to each patient’s index admission, focusing on recent discharge summaries, primary care physician (PCP) notes, and outpatient cardiology notes. This note selection process mirrors the standard chart review clinicians perform for HF patients, ensuring the inclusion of clinically relevant and temporally proximal data. Using domain expertise, we designed HF-specific bundles (Figure 1a; Table 1) to identify the critical details essential for decision-making in heart failure cases. These bundles guide the extraction process, ensuring that the summaries focus on clinically relevant information for understanding the patient’s longitudinal HF history^26^.

**Table 1.** Clinical bundle for Heart Failure Summaries. This table presents the clinical bundle for heart failure developed using domain expertise. The first column lists the category of information relevant to clinical care, the second column provides the names of the data elements within each category, and the third column offers descriptions of each element.

| Category | Heart Failure Clinical Element | Description |
| --- | --- | --- |
| Heart Failure Class and Etiology | Type of heart failure | Classification as systolic or diastolic heart failure based on cardiac function |
|  | New York Heart Association (NYHA) class | Symptom-based classification of heart failure severity and functional limitation |
|  | Type of cardiomyopathy | Identification of ischemic vs. non-ischemic etiology |
| Relevant Comorbidities | History of arrhythmias | Presence of atrial or ventricular arrhythmias |
|  | History of valvular disease | Presence of mitral or aortic regurgitation or stenosis |
| Procedures | History of ICD or biventricular pacing | Documentation of Implantable Cardioverter-Defibrillator (ICD) placement or cardiac resynchronization therapy |
|  | History of cardiac procedures | Prior interventions such as coronary angiography, stent placement, or coronary artery bypass grafting (CABG) |
| Labs / Vitals | Most recent dry-weight | The patient's weight in a euvolemic state, used to assess volume status and guide diuretic therapy |
|  | Baseline creatinine (Cr) and BNP | Laboratory markers of renal and cardiac function |
| Imaging | Recent echocardiogram findings | Ejection fraction, valvular abnormalities, thrombus presence, and structural abnormalities. |
| Medications | Goal-Directed Medical Treatment (GDMT) | Name, dosage, and frequency of GDMT medications (e.g., beta-blockers, ACE inhibitors/ARBs, aldosterone antagonists, SGLT2 inhibitors) |
|  | Diuretics | Name, dosage, and frequency of loop or thiazide diuretics (e.g., furosemide, bumetanide) |
|  | Other active cardiology medications | Name, dosage, and frequency of additional cardiac medications (e.g., nitrates, hydralazine) |
|  | Side effects and contraindications | Documented reasons for omission of cardiac medications (e.g., allergies, ACE inhibitor-induced renal failure) |
| Patient Care Team | Name of the outpatient cardiologist | Provider managing the patient's longitudinal heart failure care |
|  | Name of the outpatient electrophysiologist | Specialist overseeing arrhythmia management |
|  | Name of the primary care provider | Provider that the patient follows longitudinally for primary care |
| Disease Trajectory and Care Plan | Recent precipitating events | Documented factors contributing to recent heart failure decompensation, such as medication nonadherence or infections |
|  | Heart Failure trajectory | Assessment of whether the patient's condition is stable, improving, or deteriorating over time |
|  | Goals of care / advanced directives | Documentation of patient preferences for care escalation in the setting of progressive heart failure |

Our two-step summarization process began with the extraction of essential clinical information relevant to HF management. The HF-specific bundles served as guides for LLMs to extract pertinent information from the selected notes (Figure 1b). Extracted data were organized into the predefined bundle structure to collate information across notes (e.g., grouping ejection fraction values with corresponding timestamps). In the second step, abstraction, we leverage an LLM to synthesize the extracted information into a cohesive, readable summary, highlighting temporal trends and surfacing inconsistencies that appeared across the notes. To ensure verifiability, references—including note identifiers, note types, and encounter dates—were appended to the bottom of the summary, enabling clinicians to verify summary content against the original source material easily.

An example of a generated summary is provided in Supplemental Note 1. The structured output aligns with the HF-specific bundle, with sections corresponding to each component of the bundle. References at the end of the summary provide clinicians with direct access to the original source material, allowing them to verify the summarized information efficiently.

### Study Design for Evaluating LLM Summaries for Chart Review

We conducted in-lab evaluations with eight physicians (75% attending physicians and 25% fellows; 62.5% internal medicine, 12.5% cardiology, and 25% other specialties) to assess the impact of LLM-generated summaries on chart review efficiency and accuracy compared to standard EHR workflows. Physicians were tasked with completing a 19-item questionnaire about a patient’s heart failure history under two conditions: (1) using the EHR alone (standard chart review) or (2) using the LLM-generated summary alongside the EHR, where physicians could refer to the EHR if information in the summary were incomplete or unreliable (Figure 2a). Each physician reviewed the same eight patients, randomly sampled from our HF cohort, with the order of patients and the assigned condition (EHR Only vs. Summary+EHR) alternated to mitigate learning effects and account for potential improvements in chart review efficiency over time (Figure 2b).

**Figure 2:**
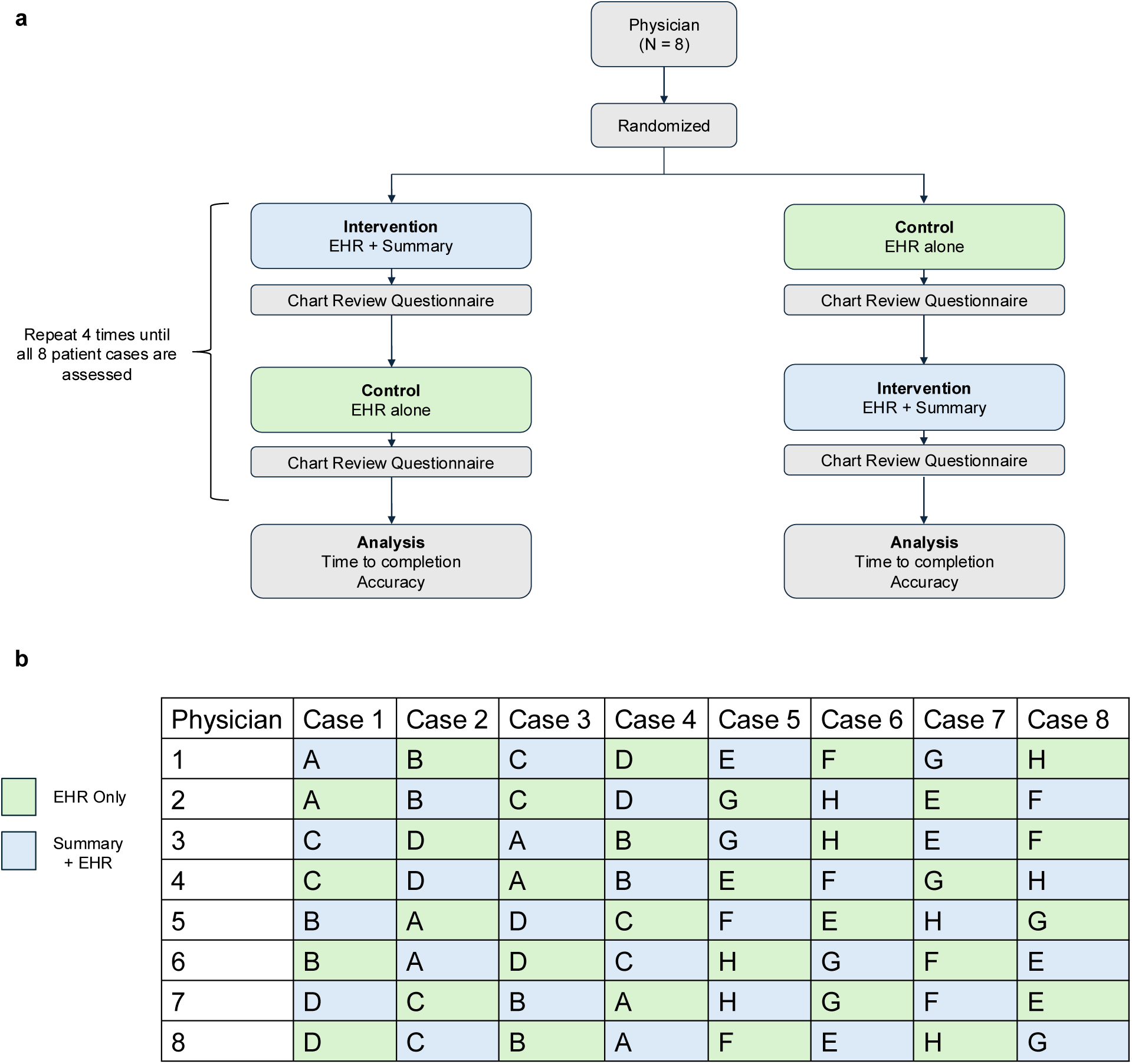
Design of the extrinsic evaluation study. (a) Each physician reviewed eight patient cases, with four cases assigned to the control condition (green), where only the Electronic Health Record (EHR) was available, and four cases assigned to the intervention condition (blue), where physicians had access to both the EHR and an LLM-generated summary to complete the questionnaire. The primary quantitative metrics were questionnaire completion time and accuracy. (b) The same eight cases, randomly selected from the heart failure cohort, were presented to all physicians. To control for order effects, the sequence in which each physician reviewed patient charts (A through H) was randomized. The cases were presented to each physician under two conditions alternately: EHR Only versus Summary + EHR.

### Impact of LLM Summary Access on Chart Review Efficiency and Accuracy

Access to LLM-generated summaries did not significantly reduce overall questionnaire completion time compared to standard chart review (mean [SD]: 9.45 [3.58] vs. 10.30 [3.95] minutes; p = 0.46, Mann-Whitney U test; Figure 3a). However, further analysis revealed variability in the impact of summary access across physicians (Figure 3b) and patient cases (Supplemental Figure 1), with some physicians and cases benefitting substantially from summary access. A multivariable regression model controlling for physician identity, case order, and their interactions explained 81.0% of the variance in questionnaire completion time (R² = 0.810, adjusted R² = 0.731; F(17,41) = 10.26, p < 0.001). Physician identity (p < 0.001) and case order (p < 0.001) were significant predictors, with physicians becoming faster as they progressed through the questionnaire —averaging 12.50 minutes for their first case compared to 8.57 minutes for the last case. While summary access alone was not a significant predictor (p = 0.18), its interaction with physician identity was (p = 0.015), suggesting individual differences in how summaries supported workflow.

**Figure 3:**
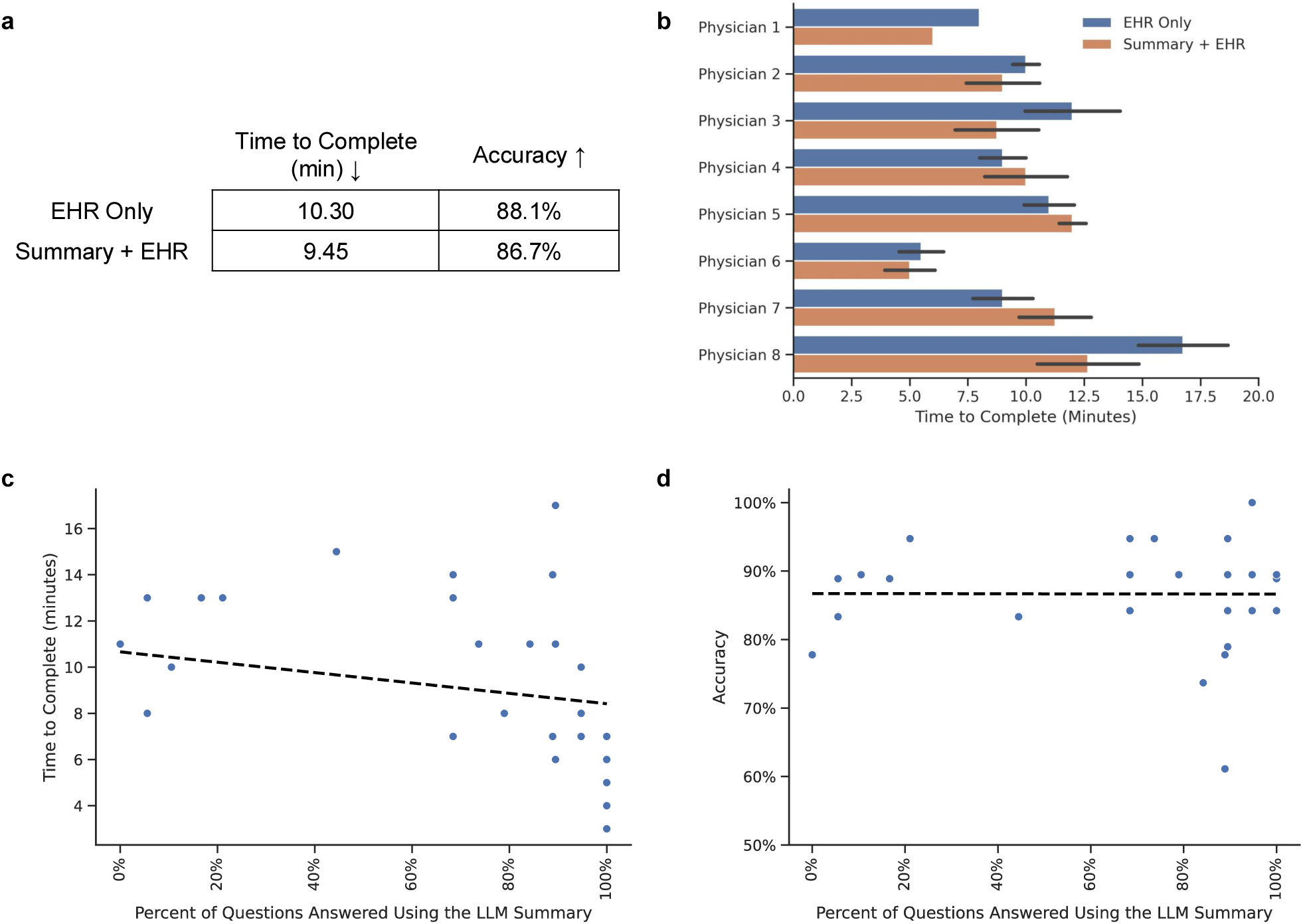
Impact of LLM-generated summaries on questionnaire response time and accuracy. (a) Overall time to completion and accuracy across both study arms. (b) Questionnaire completion times for individual physicians using the EHR Only (blue) versus EHR with LLM summaries (orange). (cd) Relationship between summary use and (c) time to completion and (d) accuracy within the Summary + EHR arm; each point represents a patient case. A linear regression line with 95% confidence interval is shown to illustrate the direction and strength of the association. See Supplemental Figure 3 for results across both study arms.

Accuracy was unaffected by summary access. The Summary+EHR and EHR-only groups had comparable accuracy (mean [SD]: 86.7% [7.7%] vs. 88.1% [8.0%]; p = 0.42, Mann-Whitney U test; Figure 3a). A multivariable regression model explained only 37.9% of the variance in accuracy (R² = 0.379), and the overall model was not statistically significant (F(17,41) = 1.474, p = 0.153). Summary access (β = 0.0134, p = 0.838) and its interactions with physician identity or case order were also non-significant.

### Impact of LLM Summary Usage on Chart Review Efficiency and Accuracy

When provided access to the summary, physicians chose to rely on it exclusively for 69.5% of questions, highlighting its perceived utility. Usage was highest for topics such as cardiac procedures, dry weight, and valvular disease (82.7% for each) and lowest for contraindications for ACE inhibitors (27.6%) and diuretic dosing (51.7%; Supplemental Figure 2).

While summary access did not significantly improve chart review efficiency for all physicians, greater summary use was strongly correlated with faster questionnaire completion (Spearman’s ρ = -0.62; p = 0.0003; Figure 3c, Supplemental Figure 3a). To further explore this relationship, we modeled completion time as a function of the percentage of questions each physician answered using the summary. Controlling for physician identity, case order, and their interactions, the model explained 86.1% of the variance in completion time (R² = 0.861, adjusted R² = 0.804; F(17,41) = 15.00, p < 0.001). An ANOVA revealed that summary usage significantly contributed to explaining variation in completion time (p = 0.002), and this effect varied significantly across physicians (p < 0.001). Physician identity (p < 0.001) and case order (p < 0.001) were also significant predictors. These findings highlight substantial variability across physicians in how effectively they leveraged the summary, suggesting that its impact depends on individual physician workflows and preferences.

Furthermore, summary use did not affect accuracy (Spearman’s ρ = 0.137; p = 0.476; Figure 3d; Supplemental Figure 3b). When physicians elected to use the summary, their accuracy was comparable to that of relying solely on the EHR (88.0% vs 86.4%, respectively; p = 0.51, chi-squared test).

### Physician Feedback on LLM Summaries

Upon completing each questionnaire using a summary, physicians rated the summaries on clinical utility, comprehensiveness, organization, relevance, and ability to surface ambiguities or inconsistencies. After completing all patient questionnaires, physicians provided overall feedback on their experience, including their likelihood of using the summaries in clinical practice, their perception of the summaries’ potential to save time, and free-text comments highlighting strengths, weaknesses, and suggestions for improvement.

All physicians indicated they were “likely” or “very likely” to use the summaries in clinical practice, with 87.5% stating the summaries would save them time. Physicians generally rated the summaries positively across key dimensions (Figure 4). Most found that the summaries contained relevant information (65.5% summaries rated as “Very” or “Somewhat” relevant), were highly comprehensive (89.7% rated as “Very” or “Somewhat” comprehensive), and well-organized (82.8% rated as “Very” or “Somewhat” organized). Additionally, 93.1% deemed the summaries “Very” or “Somewhat” clinically useful for chart review.

**Figure 4:**
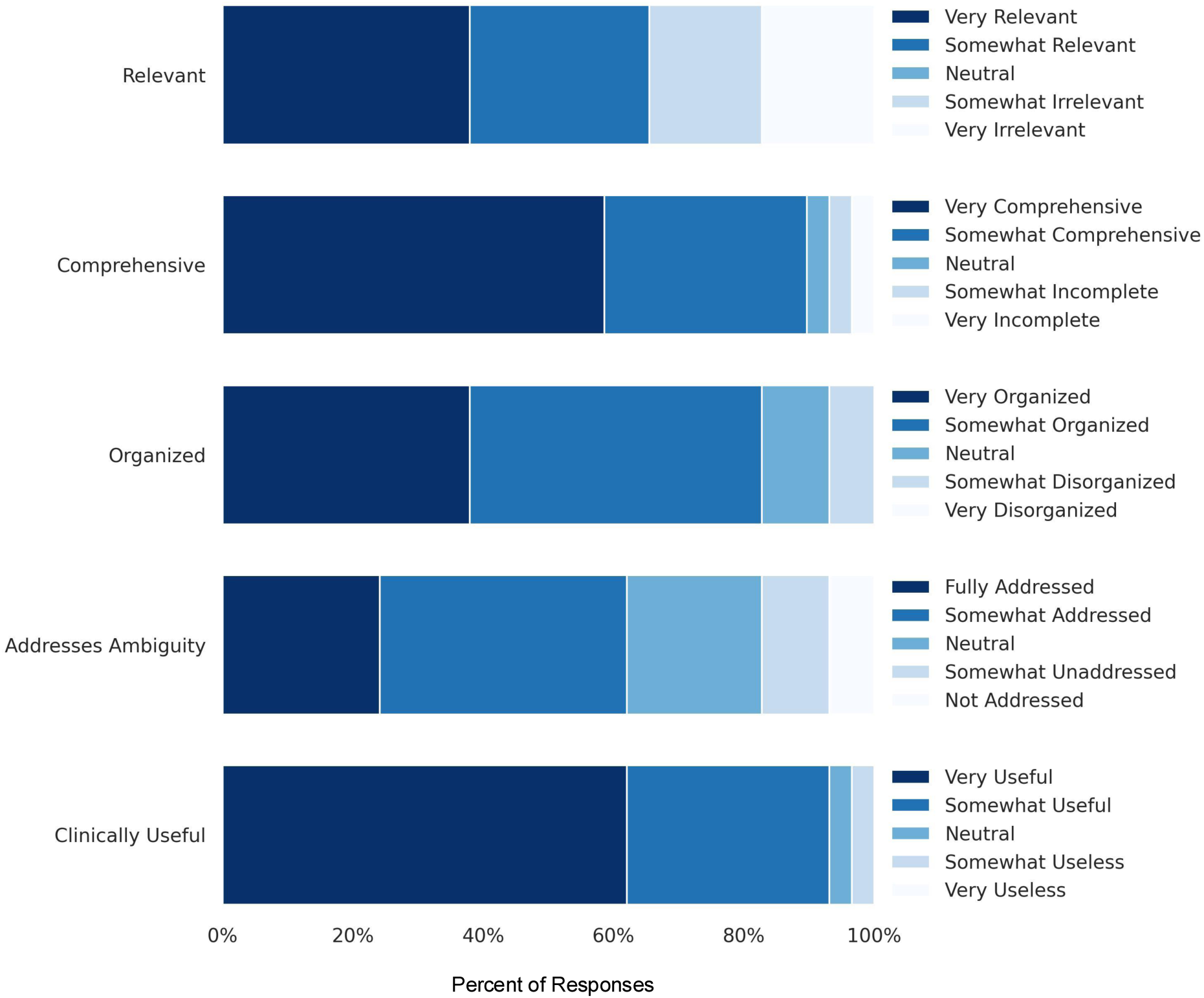
Physician assessment of LLM-generated summary quality. Distribution of Likert scale responses evaluating summary relevance, comprehensiveness, organization, ability to address ambiguity, and clinical usefulness.

However, the ability of summaries to surface ambiguous or inconsistent information received more mixed ratings, with 62.1% finding them effective but 37.9% highlighting room for improvement.

Qualitative feedback can be categorized into three main themes: strengths, limitations, and suggestions (Tables 2 and 3). Physicians praised the summaries for their organized categories and succinctness. References were especially valued for allowing verification of summary content. They found the summaries easier to navigate than EHRs. Specific information, such as medications and their related narratives, was deemed quite helpful for decision-making. Additionally, they appreciated that these summaries provided context instead of just one-word mentions. However, some physicians noted that summaries sometimes lacked essential context for the information, which led to confusion. A few found the organization counterintuitive to their thought processes. In certain instances, they preferred relying on their familiarity with EHRs to answer specific questions. There were also a few situations where physicians observed inaccuracies or missing information, which diminished their trust and limited the summaries’ use in evaluations. To enhance the summaries, physicians suggested design and layout modifications to improve usability. One such example was to implement a hover feature for the references to avoid scrolling down to the bottom of the summary each time to ascertain the source of the information. Another suggestion was to incorporate additional metadata components, such as the authorship of the information, which could offer further insight into its accuracy. Some also proposed broader uses for the summaries, such as integrating them into downstream clinical workflows.

**Table 2.** Physician perceptions of LLM-generated summaries: strengths and limitations. Themes and subthemes from the qualitative analysis, reflecting what physicians appreciated and found lacking in the summaries, along with illustrative quotes for each subtheme.

| Strengths of the summaries identified by the respondents with quotes | Limitations of the summaries identified by the respondents with quotes |
| --- | --- |
| **1. References**<br>*“I liked the fact that there were references for verification”*<br>*“However, the clinician still has to dig into the EMR to find out details like what type of arrhythmia, but it’s helpful to have the reference from the summary.”*<br>**2. Organized**<br>*“Love how pertinent data is parsed out into categories/subheadings”*<br>*“They were succinct and contain all the information in one place.”*<br>*“pulls all relevant info and arranges it in a manner that is easy to find.”*<br>**3. Useful/High yield**<br>*“The “medication narrative and changes” section is fantastic and very high yield.”*<br>*“Most of the facts that were included were quite helpful (e.g. the Cr, types of interventions). I would like to use this in real life as a great first pass on HF or to discuss with a consultant/team”*<br>*“For those that had the NYHA class, that was useful. I appreciated the medication narrative at the bottom. History of interventions was useful.”*<br>**4. Convenient/Quick**<br>*“Easier to find that having to go through EHR.”* | **1. Better familiarity with the EHR**<br>*“Summaries were also a bit long, so I sometimes went back to the EHR (which I was more familiar with) to find the data I needed”*<br>*“At times, I had to jump around to find the information I was looking for, but this could have just been related to getting used to the layout.”*<br>**2. Organization/Layout**<br>*“The organization of the summary is somewhat counterintuitive in a way that fragments my cognitive workflow a bit. My natural thought process is type/cause of heart failure f/b the most pertinent clinical data for a rapid prognostic overview of the patient’s clinical picture -- most pertinently: coronary interventions, h/o valvular dz/arrhythmias, PPM/ICD presence, meds (with recent changes), THEN the data that i can look at if i need more information/data that is more easily found in the EHR (ie proBNPs, Cr, dry weight, etc)”*<br>*“I wonder if structuring with how we structure our assessment and plan for CHF might be useful. Something like Preload, Afterload, Contractility, GDMT or something similar. “*<br>**3. Lack context**<br>*“cardiac med narrative is a little confusing - would be nice to separate into IP and OP events (or to set expectations for what is in a narrative - most recent hospitalization?, purpose/scope unclear; timing also very unclear.)”* |
| *"It will provide reassurance and a quicker way to understand some information about the patient. It highlights key things one needs to know about a heart failure patient when they walk into the door."*<br>*"Easy to find what should be structured data but is not inputted that way in notes (ie dry weight, history of arrhythmia) to answer simple questions"*<br>**5. Contextual information was present**<br>*"There are some narratives that gives context and not just one-word answers"*<br>*"provides a narrative for explanation re: med changes. Summarizes important studies/procedures."* | *"in real life - comorbidities may be more relevant (e.g. if they have bleeding issues etc.) or they may have multiple medical problems besides cardiac that complicate their management - the scope was fine for this research but in real life people may want more"*<br>**4. Accuracy**<br>*"I was primarily concerned with accuracy of information. There was one summary that was completely wrong in terms of the factual data contained. This finding made me concerned about trusting the summary in general."*<br>*"At times there were items that were not accurate and needed confirmation. It would be good to know what is confirmed and by whom."*<br>**5. Missingness**<br>*"all the fields that just said "NA" (such as for type of heart failure when the patient did actually have some HF). Then I stopped using the summary as much for that patient and just relied on the EMR"*<br>*"the narratives leave out some meds; sotimes [sic] med list includes more than one med of the same type (confusing). it doesn't include date med was last dispensed."*<br>*"Sometimes it did not contain all the info I would have wanted"*<br>**6. Redundant**<br>*"Unorganized Occasionally redundant"* |

**Table 3.**
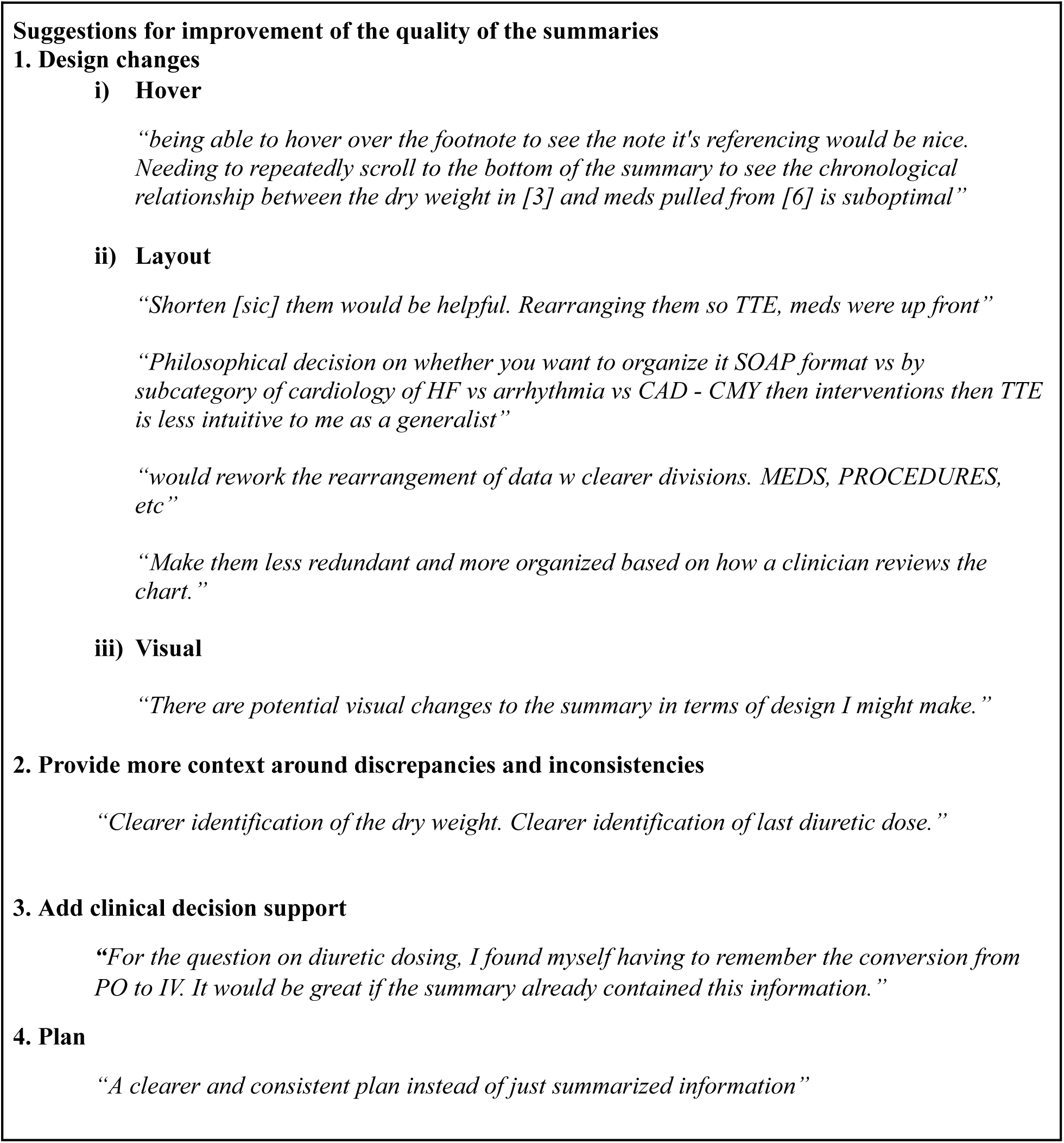
Physician suggestions for improving LLM-generated summaries. Themes and subthemes capturing physician feedback on ways to enhance summary quality, supported by representative quotes for each subtheme.

## Discussion

LLMs hold significant promise in generating clinically relevant summaries that help mitigate the cognitive burden of synthesizing longitudinal patient records. In this study, we developed and evaluated an LLM-based approach to generate problem-based admission summaries of longitudinal patient records to facilitate chart review. Using disease-specific bundles within an extract-then-abstract framework, we generated verifiable summaries for patients with heart failure that (1) prioritized clinically relevant information for disease management and (2) linked key details back to source documents. Our pilot study assessing the utility of these summaries for chart review demonstrated that physicians who relied on them frequently completed chart review tasks more efficiently, without compromising accuracy. Physicians found the summaries well-organized, comprehensive, and clinically useful, with most indicating they would integrate them into practice. Importantly, our structured approach to summary generation—guided by disease-specific bundles—provides a scalable methodology that can be adapted to other medical conditions and workflows.

We reconceptualize summarization as a series of question-answering (QA) tasks defined by disease-specific summary bundles. Unlike general summarization approaches, which risk including extraneous details or omitting critical medical information, our method structures the process around predefined bundle elements, ensuring alignment with clinical decision-making needs. This modular approach allows individual components of the bundle to be improved and evaluated separately and repurposed across different diseases by leveraging a library of common, clinically relevant elements. Importantly, this does not imply that summarization tools should be replaced by interactive QA systems alone. Predefined summaries reduce cognitive overhead by proactively extracting and organizing the information clinicians need, eliminating the burden of recalling which details are most relevant. By doing so, we take advantage of the “checklist effect”, the phenomenon where the mere presence and use of checklists can lead to improved safety and reduced errors by standardizing processes, enhancing communication, and decreasing reliance on memory^29–31^. Furthermore, unlike LLM-generated clinical notes, which may interfere with how clinicians synthesize information^32^, the summaries are designed to support rather than replace clinical reasoning. By surfacing structured, relevant information—including potentially ambiguous or inconsistent details across the chart—they preserve clinicians’ ability to reflect, interpret, and document in their own words.

One of the most critical challenges in deploying LLMs for clinical summarization is the risk of hallucinations, where models generate incorrect or misleading content^33,34^. To address this, our framework incorporates references linking summary content to its source notes, allowing clinicians to verify information quickly. Qualitative feedback highlighted that physicians appreciated this feature, emphasizing the need for transparency in AI-generated outputs. Several physicians noted that having references made verification easier, particularly when evaluating inconsistencies across notes in patient records. This aligns with previous research emphasizing that trust in AI-assisted decision-making depends on the ability to verify system outputs^35,36^. Future work should refine these verification mechanisms, integrating user interface enhancements that allow clinicians to seamlessly navigate between summaries and source notes.

Most evaluations of generative AI in healthcare focus on intrinsic measures of generated text quality rather than real-world usability^37–41^. To our knowledge, this is the first study to conduct an extrinsic physician-based usability evaluation of LLM-generated clinical summaries. As noted by Sittig and Singh^42^, new technologies must be evaluated in the social environments where they will be implemented, as clinicians will not adopt tools that fail to reduce cognitive burden or save time. Our findings reinforce this perspective: while accuracy was maintained, physicians exhibited variability in how effectively they leveraged the summaries, highlighting the importance of user-centric evaluations. Notably, while summary access did not significantly reduce overall questionnaire completion time, physicians who relied more heavily on the summaries completed questionnaires faster. We expect these results to represent an upper bound on questionnaire completion time, as clinicians may become faster with increased familiarity with the summary layout. Usage patterns varied across question types—summaries were most useful for confirming the presence of clinical information (e.g., cardiac procedures, dry weight) but were less relied upon for tasks requiring identification of absent information (e.g., ACE-inhibitor contraindications). This may reflect that it is cognitively easier to verify the presence of information in a summary than to trust that its absence means the information is truly missing from the record. These findings suggest that AI-generated summaries may be particularly valuable for synthesizing confirmatory information rather than surfacing negative findings, an insight that could inform future improvements in clinical summarization tools.

One possible explanation for the limited improvement in questionnaire completion time with summary access is the presence of algorithm aversion or selective adherence bias^43,44^. Although we did not directly measure this in our study, qualitative feedback suggests that even minor perceived inaccuracies led some physicians to disregard the summaries entirely, limiting their overall utility. Future work should explore how different clinician subgroups perceive and tolerate errors, with the goal of establishing thresholds for acceptable accuracy and designing strategies to mitigate uncertainty in AI-generated outputs.

Despite the lack of statistically significant time savings, all of the physicians said they were likely or very likely to use this in clinical practice, with 87.5% stating it would improve speed. This disconnect may reflect the limitations of the questionnaire-based evaluation, which likely underestimates the value of the summaries. In real-world settings, clinicians must recall relevant questions each time they review a chart, a process complicated by frequent interruptions, task switching^5^, and evolving information needs. Even in uninterrupted settings, chart review is rarely a one-time task; clinicians revisit patient records at multiple decision points, such as before admission, during documentation, when making treatment decisions, and before rounds. Physicians may have considered the broader utility of the summaries across these more complex, cognitively demanding scenarios—where having a structured, problem-based summary could be especially valuable.

The extrinsic evaluation further revealed that physicians with access to the full EHR achieved only 88% accuracy in answering the questionnaire questions. This highlights the inherent difficulty of synthesizing information from longitudinal records, which often contain outdated or duplicated information, and underscores that the goal of AI methods should be to improve on current clinical workflows rather than achieve perfect performance. It also suggests the importance of user studies to better understand what constitutes a practical gold standard for clinical accuracy.

Physicians also demonstrated substantial variability in their preferences for the format and organization of the summaries. Some found them well-structured and easy to navigate, while others found the layout counterintuitive. These differences suggest that a universal solution may not be feasible, reinforcing the need for extrinsic evaluations to tailor summary formats to different clinical workflows. Our bundle-based approach offers a flexible way to accommodate these differences—by structuring summaries around predefined clinical elements, which can be adapted to align more closely with individual preferences. Clinicians could prespecify a template that defines which bundle components to include and how to organize them, improving usability and integration into diverse workflows.

This study has several limitations. First, we leveraged domain knowledge to define a subset of recent clinical notes for summarization. While this mirrors real-world chart review workflows, this approach may exclude older, yet clinically relevant longitudinal information. Future work should explore strategies that incorporate retrieval augmented generation or allow clinicians to specify which notes to include. Second, our framework currently focuses on unstructured text. Integrating structured EHR data (e.g., lab values, vital signs) and information from external health systems (e.g., via CareEverywhere) would enable more comprehensive summaries. Third, although this study focused on heart failure admission chart review, the methodology is broadly applicable to other diseases and clinical contexts, such as inpatient rounding, emergency department admissions, or outpatient care. Expanding to diverse conditions and workflows is necessary to assess generalizability. Fourth, this pilot was conducted at a single healthcare system using a single EHR and in a controlled environment. The limited sample size may have reduced our ability to detect statistically significant differences in efficiency outcomes, despite qualitative feedback suggesting perceived time savings. Larger, pragmatic trials embedded in real-world settings and multiple centers are needed to evaluate long-term adoption, safety, and clinical impact. Fifth, future work should refine the user interface to enhance usability—for example, by adding clickable references that link summary content to source notes within the EHR. Sixth, our goal was to develop a framework for verifiable summarization, rather than benchmarking different LLMs. Future research should apply this approach to state-of-the-art language models as they evolve. Finally, while our study focused on summarization to support chart review, AI-generated summaries hold promise for broader clinical applications, including decision support and quality improvement.

Our findings suggest that problem-based longitudinal summarization has the potential to improve chart review efficiency and standardize information presentation in clinical workflows. However, variability in summary preferences and clinician adoption highlights the need for extrinsic, user-centered evaluations to guide AI summary development. Moving forward, enabling clinician customization, integrating verifiable summarization into EHR systems, and conducting large-scale studies in real-world clinical settings will be critical to ensuring that AI-generated summaries meaningfully enhance clinical practice.

## Methods

### Patient selection

Our population included patients seen at the Mass General Brigham (MGB) system. MGB is a healthcare system containing tertiary and community hospitals and outpatient practices across New England. MGB uses the Epic electronic health record system. While the resulting methods can be readily extended to other diseases, we focused on patients with heart failure, which is a complex, chronic medical condition leading to over a million hospitalizations per year^45^. We leveraged MGB’s Research Patient Data Registry (RPDR) to retrieve the data for all patients with at least two heart failure ICD (International Classification of Diseases) codes (ICD10:I50.XX) or DRG (Diagnosis Related Groups) codes (APR v30, AP v21, MS v24) who were seen at Brigham and Women’s Hospital and Massachusetts General Hospital from 01/01/2017 to 12/19/2023. Patients with heart failure with both preserved and reduced ejection fraction are included in this cohort. We defined index admission as the most recent admission with an H&P note and a heart failure ICD or DRG code. We excluded all patients without an H&P note, those who were part of the home hospital program, and those who were transferred from an outside hospital. To ensure our cohort included patients with a relevant longitudinal history of HF, we further restricted the sample to those with at least five total clinical notes and at least one HF diagnosis code recorded before the index admission. This criterion helped exclude patients who were newly diagnosed with heart failure during the index admission. The resulting cohort included patients seen at both Mass General Hospital and Brigham and Women’s Hospital. The Institutional Review Board at Mass General Brigham approved this study.

### Development of Disease-Specific Summary Bundles

Clinicians rely on structured patterns of grouping information—known as bundles—to streamline decision-making and reduce cognitive burden in clinical practice^26^. These bundles are context-specific and vary by use case, such as inpatient rounding, outpatient visits, or addressing a particular clinical problem^26^. By structuring patient data in this way, bundles ensure that the minimal information required for decision-making is readily accessible at the point of care.

Building on this concept, we developed a heart failure-specific summary bundle (Table 2), outlining key clinical data elements essential for managing hospitalized HF patients. The bundle can be denoted as:

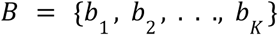

where each bundle element *b*_*k*_ represents a clinically relevant component of heart failure management, such as ejection fraction, diuretic use, or history of cardiac procedures. This bundle was designed using domain expertise to prioritize clinically relevant details, including cardiac history, prior interventions, pertinent labs, and medication use.

To generate clinically meaningful HF summaries, we structured our extraction process around these predefined bundles. This method ensures that summaries align with clinicians’ cognitive workflows and contain the information most relevant to HF management. The bundle-based approach is inherently scalable to other diseases by modifying domain-specific data elements to fit different clinical contexts.

### Generation of Verifiable Admission Summaries

We developed a structured extract-then-abstract pipeline to generate verifiable, problem-based admission summaries for patients with heart failure. This approach ensures that all extracted information is clinically relevant, traceable to source notes, and organized within a structured summary format that maintains the predefined bundle structure. The summarization process consists of four key steps: selecting relevant clinical notes, extracting disease-specific information, aggregating extracted information, and synthesizing the extracted content into a structured summary while maintaining source references.

#### Salient Note Selection

For each patient, we identified an index admission, denoted as *i*, and retrieved the set of clinical notes from encounters that occurred before this admission. The set of notes is represented as:

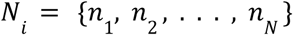

where each note *n*_*j*_ consists of a sequence of words. To ensure clinically meaningful summarization, we selected high-yield clinical notes from patient encounters before the index admission. Based on domain knowledge, we included the most recent discharge summary, history and physical (H&P) note, outpatient visit note, and echocardiogram report. We excluded Care Everywhere notes—documents shared from external institutions via Epic^46^. Emergency department (ED) notes were excluded, but same-day outpatient visit notes were retained to capture last-minute pre-admission clinical updates. Additionally, for this pilot study, we excluded structured data elements, such as lab results and vital signs, that were not embedded within clinical notes.

#### Extraction of Bundle Elements

For each salient note in a patient’s record, we applied an extraction function that extracts the relevant details for each bundle element:

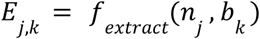

where *E*_*j*,*k*_ represents the extracted information from a note *n*_*j*_ that corresponds to a bundle element *b*_*k*_ . The extraction process was performed using GPT-4o version 5-13-2024. The system prompt used for extraction can be found in Supplemental Note 2. Each note was included in the user prompt. We set the model’s temperature to 0, top p to 1, and maximum number of output tokens to 4096. The models were accessed using Azure OpenAI in Mass General Brigham’s HIPAA-compliant Azure tenant behind the hospital firewall.

#### Aggregation of Extracted Information

Once information was extracted from individual notes, we aggregated it across all notes while preserving details about when and where each piece of information was documented. This aggregation process ensures that all available clinical information is grouped appropriately, facilitating an accurate and comprehensive summary. The extracted data was structured as a dictionary, where each entry is indexed by a tuple containing the note type and note date:

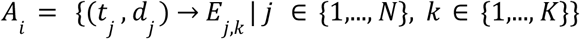

where *t*_*j*_ represents the type of note (such as a discharge summary or outpatient visit) and *d*_*j*_ is the date the note was written. The value *E*_*j*,*k*_ contains the extracted heart failure-related information for the bundle element *b*_*k*_.

#### Structured Summarization

We synthesized the extracted information into a structured admission summary that follows the bundle format. The summarization function:

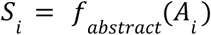

produces a structured summary, where each element in the output corresponds to a bundle element in *B*. The summarization process was performed using GPT-4o version 5-13-2024 using the same Azure infrastructure and hyperparameters described above. The model is instructed to identify and highlight inconsistencies where multiple notes reported conflicting information and include references to the original notes by appending metadata (note type and date), allowing clinicians to verify details in the EHR quickly. The system prompt used for summarization can be found in Supplemental Note 2.

#### Prompt Development

To construct a prompt development dataset, we randomly sampled eight patients from our heart failure cohort who were admitted to Brigham and Women’s Hospital (BWH). Patients from BWH were used exclusively for prompt development, while those from Massachusetts General Hospital (MGH) were reserved for physician evaluation to ensure that the evaluation was conducted on an independent dataset. For each patient in the prompt development dataset, we constructed gold-standard summaries to serve as reference outputs for evaluating extraction and summarization quality. These gold-standard summaries included two key components: (1) structured annotations detailing the specific notes and sections from which each bundle element was extracted, and (2) a final expert-curated summary following the predefined bundle structure. These references provided a benchmark for assessing the accuracy, completeness, and organization of the LLM-generated outputs.

We iteratively refined the extraction and summarization prompts by comparing model-generated outputs to these gold standards and incorporating qualitative feedback from physicians. Adjustments were made to improve the precision of extracted content, ensure alignment with clinical reasoning, and enhance the readability and usability of the final summaries within real-world clinical workflows.

### Overview of Physician Evaluation

We conducted an extrinsic evaluation of LLM-generated summaries to systematically assess their impact on clinical decision-making in real-world chart review scenarios. Using a mixed-methods approach, we integrated quantitative evaluation to measure how summaries influenced physician performance and qualitative assessments to capture physicians’ perceptions of their utility in practice. For the quantitative evaluation, we conducted a randomized crossover experimental study (Figure 2a) to compare physician performance in answering questions about a patient’s chart when using LLM-generated summaries alongside the EHR versus standard EHR-based chart review alone. Physicians were recruited using a snowball sampling methodology, leading to the identification of eight physicians from Brigham and Women’s Hospital and Massachusetts General Hospital across multiple specialties, including Internal Medicine, Cardiology, Clinical Informatics, Gastroenterology, and Allergy & Immunology. We constructed an evaluation dataset by randomly sampling eight patients from the heart failure cohort who were admitted to Massachusetts General Hospital. These cases were distinct from those used in prompt development, ensuring that model evaluation was conducted on unseen data.

### Questionnaire and Survey Design

To simulate real-world chart review, we developed a 19-question questionnaire designed to assess physicians’ ability to retrieve and synthesize salient information from a patient’s chart. These questions prompt the physician to assess the patient’s heart failure severity, comorbidities, test results, medications, and other information related to management of the patient’s heart failure (e.g., “Has this patient had an echocardiogram within the last 6 months (i.e. since March 1 2023)?”). A complete list of questions is provided in the supplementary materials (Supplemental Note 3). Physicians completed the questionnaire for each of the eight patients in the evaluation cohort. The questionnaire required participants to extract key details related to the patient’s heart failure history, mirroring the cognitive tasks performed in clinical practice. Two physicians constructed the initial questionnaire, ensuring it reflected clinically relevant information retrieval and decision-making tasks. To establish content validity, a separate researcher and physician independently reviewed the questionnaire, and their feedback was incorporated through an iterative refinement process. These refinements aimed to eliminate ambiguity, enhance clarity, and account for variability in clinical judgment, ensuring that questions were interpretable and clinically meaningful. For each question within the questionnaire, physicians were instructed to indicate the source utilized to formulate their responses. They were directed to specify “EHR” if they referenced the Electronic Health Record at any point for the verification of the information presented in the summary.

To assess physician perceptions of the LLM-generated summaries, we designed two additional surveys focused on judgment-based evaluations. The first survey, administered immediately after each questionnaire was completed with the summary, asked physicians to rate the quality of the summary on a 5-point Likert scale across five key dimensions: relevance, comprehensiveness, organization, ability to address clinical ambiguity, and overall clinical usefulness. The second survey, administered after physicians completed all patient cases, evaluated their overall impressions of the summaries. Physicians were asked to rate how likely they would be to use the summaries in their clinical practice and whether they believed the summaries would save time. Additionally, the survey included free-text questions, allowing participants to provide qualitative feedback on the strengths and limitations of the summaries, as well as suggestions for improvement. All questionnaires and surveys were designed and administered using REDCap.

### Study Protocol

Evaluations were conducted remotely via Teams or Zoom using a randomized crossover experimental design, adapted from Hirsch et al.’s evaluation of the HARVEST tool^7^. Physicians reviewed eight patient cases, completing a 19-question questionnaire per case using either (a) the EHR only or (b) an LLM-generated summary of the patient’s heart failure history + EHR (Figure 2a). To control for physician variability and case difficulty, physicians alternated between conditions, and cases were presented in eight different sequences to minimize learning effects (Figure 2b).

Before starting the evaluation, physicians received written and verbal instructions outlining the study protocol (Supplemental Note 4). They were also guided on configuring EHR filters to restrict access to notes documented before the index admission date at Mass General Brigham (*i.e.,* excluding Care Everywhere records).

Physicians were instructed to complete the questionnaire as accurately and efficiently as they would during real-world chart review. They were allowed to reference any pre-admission data available in the EHR. To measure task efficiency, we recorded the time required to complete each questionnaire, with a maximum time limit of 30 minutes per case. This limit was determined based on pilot testing and was deemed sufficient for completing the questionnaire under realistic clinical conditions. If the time limit was reached, the survey automatically closed.

### Statistical Analysis

We compared physician performance between the EHR-only and Summary+EHR conditions using two primary metrics: time to completion and accuracy. We hypothesized that automatically generated summaries would reduce task completion time without compromising accuracy in answering clinical questions.

Time to completion was measured through physician-recorded start and end times for each questionnaire. Accuracy was evaluated by comparing each physician’s responses to a gold-standard answer set. To construct the gold-standard responses for each patient, two team members—one with a clinical background and one with a research background—conducted a detailed, time-unconstrained chart review to ensure comprehensive and precise reference answers. To maintain consistency in free-text responses, all physician-submitted answers were standardized to match the gold-standard format. Dry weight values were converted to kilograms, and diuretic doses were normalized to equivalent furosemide IV doses.

When multiple dry weight values were clinically reasonable, the gold standard included a range or set of valid values, with responses outside this range classified as incorrect. Similarly, the gold-standard diuretic dose specified a minimum effective dose (relative to the patient’s home dose), and any response meeting or exceeding this threshold was considered correct.

We assessed the impact of LLM-generated summary access on chart review efficiency and accuracy using a combination of two-sided non-parametric tests, multivariable regression models, and correlation analyses. To model questionnaire completion time and accuracy, we fit ordinary least squares (OLS) regression models with summary use, physician identity, case order, and their interactions as predictors.

Summary use was operationalized either as a binary variable (Summary+EHR vs. EHR alone) or as a continuous measure (percent of questions answered using the summary), depending on the model. Model selection was guided by the Akaike Information Criterion (AIC), which balances model fit and complexity; including patient identity worsened model fit according to AIC and was therefore excluded. We performed type II analysis of variance (ANOVA) to quantify the contribution of each term to explained variance and assess the significance of main effects and interactions. This approach enabled us to estimate both the independent and physician-specific effects of summary access while accounting for learning effects over time. Residuals were examined to confirm OLS assumptions. Responses from the first four patient cases reviewed by Physician 1 were excluded, as these were used to refine the questionnaire during an initial phase of evaluation.

#### Task Completion Time Analysis

Given the non-normal distribution of completion times, we used a Mann-Whitney U test to compare time to task completion between the EHR-only and Summary+EHR conditions. To account for physician variability and case order effects, we constructed a multivariable regression model, including physician identity, case order, and their interactions as covariates. Model fit was assessed using R² and adjusted R², and statistical significance was determined via F-tests and p-values for individual predictors.

To evaluate the relationship between summary usage and efficiency, we calculated Spearman’s rank correlation coefficient (ρ) between the percentage of questions answered using the summary and questionnaire completion time. We further conducted a regression analysis, controlling for physician identity, case order, and their interactions, to assess whether frequent summary use influenced efficiency.

#### Accuracy Analysis

We compared physician accuracy between conditions using a Mann-Whitney U test, with accuracy defined as the proportion of correct responses per case. A multivariable regression model was used to assess whether summary access or physician-level factors predicted accuracy. Additionally, we examined whether summary usage behavior influenced accuracy using a chi-squared test to compare accuracy rates between responses derived from the summary versus the full EHR.

#### Likert Scale Analysis

We analyzed physician ratings of summary quality across five dimensions: relevance, comprehensiveness, organization, ability to address ambiguity, and clinical usefulness. For each dimension, we calculated the proportion of responses in each Likert scale category.

All analyses were conducted using Python 3.8.0 using the pandas, numpy, statsmodels and Scipy packages for regression modeling, ANOVA, and non-parametric statistical tests.

### Qualitative Analysis

Free-text responses from the final survey were analyzed to identify emerging themes and subthemes related to physician perceptions of the LLM-generated summaries. Two researchers independently coded and reviewed each response, reconciling their analyses to develop a final set of themes and subthemes. Discrepancies were resolved through discussion to ensure consistency in thematic categorization. Excel was used to facilitate data organization and analysis.

## Author Contributions

E.A., R.V., and LZ conceptualized the study. E.A. and R.V. designed the study, and E.A. secured funding for the study. R.V. and Z.S. created the heart failure bundle and the gold standard summaries. R.V., Z.S., and E.A. designed the questionnaire, and L.C. validated the questionnaire. E.A. developed the “extract-then-abstract” LLM approach and conducted all statistical analyses. R.V. and E.A. performed the qualitative analyses. L.C., K.R., E.G., C.H., M.K., J.R., D.T., O.U., J.G.Y. evaluated the LLM-generated summaries. R.V. and E.A. drafted the manuscript, which was edited and critically reviewed by Z.S., L.C., K.R., E.G., C.H., M.K., J.R., D.T., O.U., J.G.Y., L.Z., and D.B. Mass General Brigham was the source of data and location for all analyses. All authors read and approved the final manuscript and had final responsibility for the decision to submit it for publication.

## Acknowledgments

This work was supported by the Microsoft Research Accelerate Foundation Models Academic Research Program, which provided funding for access to the Azure OpenAI API. ZS was supported by the Massachusetts General Hospital ECOR Fund for Medical Discovery Clinical Research Fellowship Award and National Library of Medicine Research in Emerging Areas Critical to Human Health LRP. JR was supported by funding from the National Institute on Minority Health and Health Disparities. DT was supported by a NIH grant [T32DK135449]. JGY was supported by the National Library of Medicine/National Institutes of Health grant [T15LM007092].

## Competing Interests

EG reports equity holdings in the startups Uzobi, Inc. and Tendo, as well as stock ownership in Amgen, Eli Lilly, GlaxoSmithKline, Johnson & Johnson, and Pfizer. DB reports grants and personal fees from EarlySense, personal fees from CDI Negev, equity from Valera Health, equity from CLEW, equity from MDClone, personal fees and equity from AESOP Technology, personal fees and equity from FeelBetter, and grants from IBM Watson Health, outside the submitted work. EA reports consulting fees from Fourier Health. None of these entities had any role in the design, execution, evaluation, or writing of this manuscript. All other authors declare no competing interests.

## Data Availability

The MGB data used in this study contains identifiable protected health information and, therefore, cannot be shared publicly. MGB investigators with appropriate IRB approval can contact the authors directly regarding data access.

## Code Availability

The code used to generate summaries of clinical notes can be found at the following GitHub repository: https://github.com/EmilyAlsentzer/longitudinal_ehr_summarization.

## Notes

### Author Declarations

The Institutional Review Board of Mass General Brigham waived ethical approval for this work.

