## Supplementary Information for "Verifiable Summarization of Electronic Health Records Using Large Language Models to Support Chart Review"

### Supplementary Figures and Tables

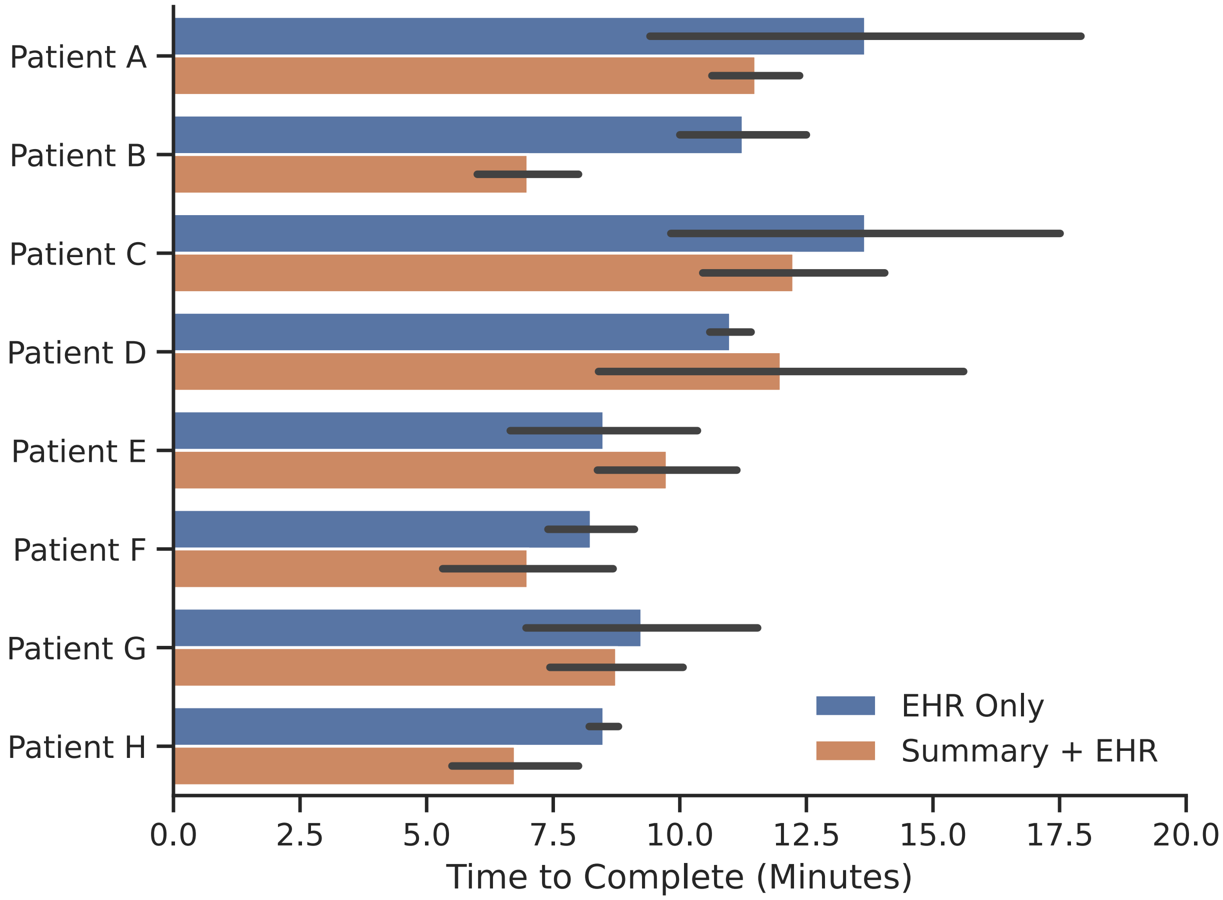


**Figure S1:** Time to complete patient case questionnaires using only the EHR (blue) compared to using LLM-generated summaries with the EHR (orange).

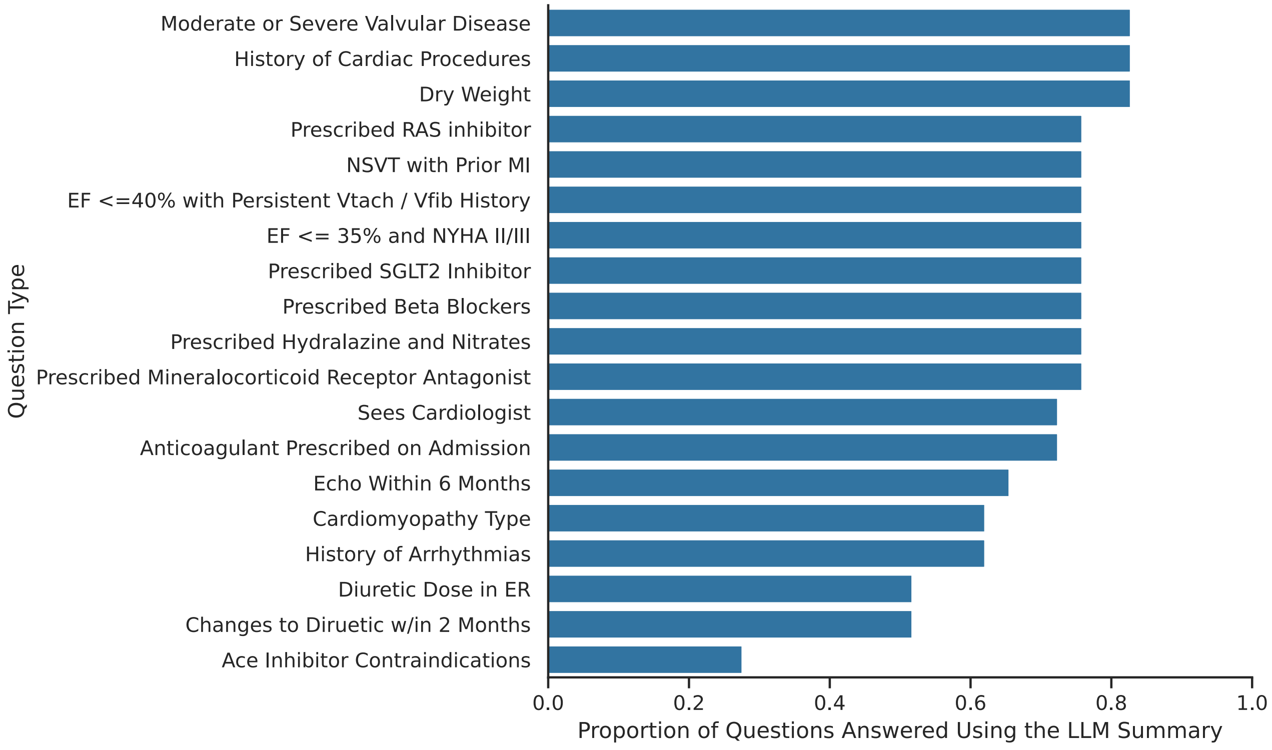


**Figure S2:** The proportion of questions that were answered using the LLM summary by question type.

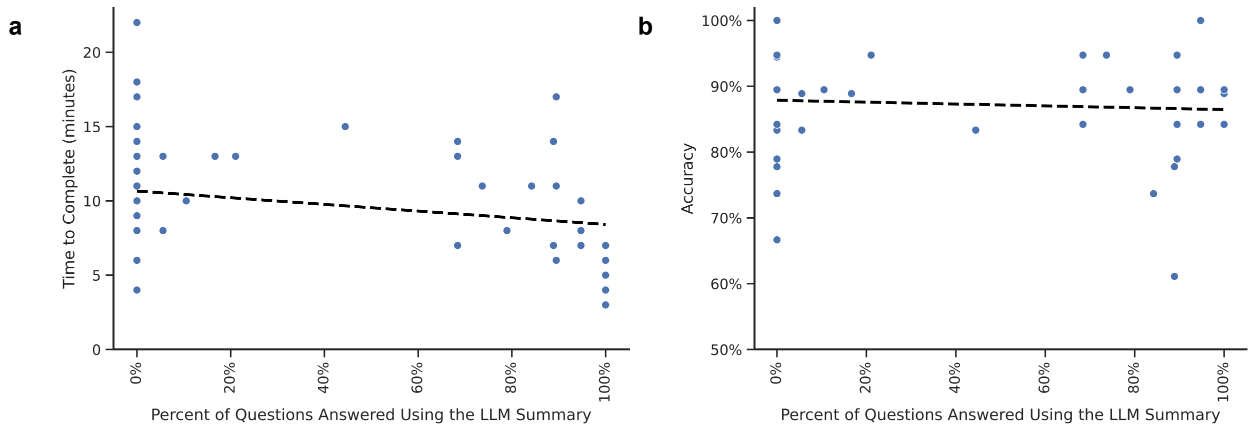


**Figure S3: Relationship between summary use and questionnaire outcomes.** (a) Time to completion and (b) accuracy, shown as a function of summary use. Each point represents a patient case. Includes responses from both the EHR-only and Summary+EHR arms; summary usage was 0% for physicians in the EHR-only arm as they didn't have access to the LLM summary. See Figure 3c–d for results limited to the Summary+EHR arm.

### Supplementary Notes

**Supplementary Note S1. Example of a Large Language Model–Generated Clinical Summary.** All protected health information (PHI) has been redacted and replaced with placeholders (e.g., [NAME], [DATE]). Clinical details have been modified to preserve the structure and format of the summary while removing any potentially identifying or unique clinical characteristics.

**Name:** [Name]
**MRN**: [MRN]
**Admission Date**: [Date]

#### Heart Failure Summary

**Type of Heart Failure**: HFrEF (Chronic systolic congestive heart failure) [1,2,4]

**NYHA Class:** NA

**Ejection Fraction**
- Most recent EF from last TTE ([DATE]) is 21% [3]
- Historical range: EF 21% consistently reported in [MONTH] [YEAR] [1,2,3]

**Dry Weight**
- Most recent lowest measurement post-discharge: 185 lbs on [DATE] (Discharge note) [1]
- Range of recorded weights: 180-200 lbs from [DATE] to [DATE] [1,2,3,4]
- Context: Significant weight loss recently, new dry weight likely in the 180s [1]

**Creatinine**
- Most recent value: 0.73 mg/dL on [DATE] (Office Visit note) [4]
- Range of values: 0.70-0.85 mg/dL from [DATE] to [DATE] [1,2,4]

**NT-proBNP / BNP**
- Most recent value: NT-proBNP 6,781 pg/mL on [DATE] (Office Visit note) [4]
- Range of values: NA

**Type of Cardiomyopathy**: Non-ischemic cardiomyopathy (NICM) [1,2,4]

**Cause of Cardiomyopathy:** NA

**History of Coronary Interventions**
- [DATE]: Cath showed clean coronaries [1]
- [DATE]: Dual-chamber ICD placement [1,2,4]
- [DATE]: ICD upgraded to CRT-D [1,2,4]

**Recent TTE Findings**
- TTE on [DATE]:

- Severely impaired LV systolic function, EF 21%
- Severely dilated LV, normal RV function, dilated LA
- Moderate functional MR, RVSP 43 mmHg [1,2,3]

**History of Valvular Disease**
- Moderate functional mitral regurgitation [1,2,3]
- Trace tricuspid and pulmonary regurgitation [3]

**History of Arrhythmias or VTach**
- History of VT, SVT, and NSVT [1,2]
- Recent episodes of SVT and wide complex tachycardia [1]

**Pacemaker or ICD Presence**
- CRT-D (upgraded from dual-chamber ICD) [1,2,4]

**Diuretic Medications and Dosage**
- Torsemide 100 mg daily (home) [2,4]
- Torsemide 30 mg daily (discharge) [1]

**Cardiac Medications and Dosage**
- Metoprolol succinate 50 mg daily [1,4]
- Sacubitril-valsartan 24-26 mg BID [1,4]
- Atorvastatin 80 mg daily [1,4]
- Aspirin 81 mg daily [1,4]
- Empagliflozin 25 mg daily [4]

**Cardiac Medication Narrative and Changes**
The patient was initially on a regimen including metoprolol succinate, sacubitril-valsartan, atorvastatin, aspirin, and torsemide. Due to food poisoning and vomiting, the patient stopped taking medications at home. During the recent hospital admission, torsemide was reduced from 100 mg to 30 mg as the patient was eating well and GI symptoms had resolved. Entresto (sacubitril-valsartan) was held until further instructions from [NAME]. The patient was also given IV magnesium sulfate, amiodarone bolus, and D50 during an episode of wide complex tachycardia. The current regimen includes metoprolol succinate 50 mg daily, sacubitril-valsartan 24-26 mg BID, atorvastatin 80 mg daily, aspirin 81 mg daily, and empagliflozin 25 mg daily [1,2,4].

**Outpatient Cardiologist(s)**: [NAME] [1,2]

**Electrophysiologist(s):** NA

**Primary Care Physician (PCP)**: [NAME] [1,3,4]

**Precipitating Events for HF**
- Stopped taking medications due to food poisoning and vomiting [1,2]

**HF Trajectory**
- Worsening, with recent admissions for HF exacerbations in [MONTH] [YEAR] and [MONTH] [YEAR] [1,2,4]

**Most Recent HF Exacerbation**
- [MONTH] [YEAR]: Admitted for HF exacerbation, also treated for COPD exacerbation and pneumonia [1]
- [MONTH] [YEAR]: Admitted for acute heart failure [2]
- [DATE]-[DATE]: Admitted for COPD and CHF exacerbation, responded well to IV diuresis, transitioned to torsemide, and started on GDMT [4]

**Comorbidities**
- [AGE]M with HFrEF (EF 21%), VT, SVT, severe COPD on 3L NC at home, T2DM on insulin, HTN, HLD, OSA, iron deficiency, gait instability, chronic pain, anxiety, depression, LBBB, and history of medication noncompliance [1,2,4]

**Advanced Directive**
- Health Care Proxy: [NAME]
- Code Status: Full Code [1]
- DNR/DNI per MOLST in [YEAR] [2]

**Other Pertinent Info**
- Discharged home in stable condition with VNA nursing [1]
- Significant weight loss of 15 kg due to poor PO intake and loss of appetite [2]

**References**
[1] Discharge Note, [DATE]
[2] H&P Note, [DATE]
[3] Echo Report, [DATE]
[4] Office Visit Note, [DATE]

**Supplementary Note S2. Prompts Used for Information Extraction and Summary Generation in the Extract-Then-Abstract Framework.**

<u>Extraction Prompt</u>

```
As an expert cardiologist at a leading medical institution, your task is to
meticulously extract heart failure-related medical history from patient notes. Use
the following guidelines to structure your response:

Data Extraction: For all relevant heart failure elements listed below, extract the
information from the patient's note. Make sure to extract the minimal but
sufficient amount of surrounding text to understand the context (e.g. relevant
dates of lab values). Do not assume or infer information not explicitly stated in
the note. You can extract multiple different sentences / phrases for each element.
```

Formatting Dates and Units: For all measurements, include appropriate units. Record the date of each measurement, considering that notes may include data from previous visits. For instance, if a note mentions a measurement from a prior discharge, include the discharge date. Present dates in the format MM/DD/YYYY.

Handling Multiple Values and Contradictions: If there are multiple values for an element, include all mentions, making sure to include pertinent dates or relative dates (e.g. "yesterday (6/11/22)", "according to H&P from 6/5/22") if mentioned.

Below are the heart failure elements to extract, with descriptions for clarity:

Type of Heart Failure: Identify whether the heart failure is systolic, diastolic, etc.
NYHA Class: Class of heart failure indicating severity.
Ejection Fraction: Ejection fraction percentage, with date; mention if value from an echo.
Dry Weight: Patient's weight without extra fluids, with most recent pertinent date; Note if it is weight on discharge or previously recorded dry weight; Extract any text describing weight or changes in weight and the context in which the weight was taken (e.g. last visit to cardiologist, on admission, on discharge)
Creatinine: Baseline Creatinine lab value, with date.
NT-proBNP / BNP: Baseline NP-proBNP or BNP lab value, with date
Type of Cardiomyopathy: Ischemic, non-ischemic, or both.
Cause of Cardiomyopathy: whether due to CAD, autoimmune disorders, etc.
History of Coronary Interventions: History of cardiac procedures, such as coronary angiography, stents, CABG, right heart cath, pacemaker/ICD placement, TAVR or other valve replacement surgeries, congential heart disease procedures, cardioversion, heart transplant, cardiac biopsies; Include pertinent results from recent angiogram if present.
Recent TTE Findings: Description of most recent and pertinent findings, including EF; Note any issues with obtaining high quality results if present; Include date of TTE.
History of Valvular Disease: Description of history of valvular disease, such as regurgitation or stenosis.
History of Arrhythmias or VTach: Description of history of Afib, Aflutter, Vtach, VT arrest, or Vfib, including pertinent interventions.
Pacemaker or ICD Presence: Details about any ICD, including whether CRT or dual chamber.
Diuretic Medications and Dosage: List diuretics with dosage and frequency that patient is on. Denote home meds vs inpatient medications
Cardiac Medications and Dosage: List medications, dosages, and frequency that patient takes; include beta blockers, ace inhibitors, ARB, ARNI, SGLT2 inhibitors, aldosterone antagonists, hydralazine, nitrates, anticoagulants if present. Differentiate between medications in the hospital vs home meds.
Cardiac Medication Narrative and Changes: All sentences related to the cardiac medication plan, including current meds, side effects, contraindications, and

recent changes in cardiac medications with reasons for the changes; extract any reasons for why the patient may/may not be on goal-directed medical therapy, if mentioned. Clarify meds taken in hospital vs at home.
Outpatient Cardiologist(s): Names of cardiologists seen in an outpatient setting. Do NOT extract the attending cardiologist in the hospital)
Electrophysiologist(s). Names of electrophysiologists seen in an outpatient setting.
Primary Care Physician (PCP): Patient's outpatient PCP.
Precipitating Events: Known reasons for worsening HF, if present.
HF Trajectory: Overall disease trajectory (stable or worsening).
Most Recent HF Exacerbation: Date and description of the most recent hospitalization for HF exacerbation.
Comorbidities: Other relevant/significant comorbidities that the patient has. Make sure to extract the "one-liner" for the patient.
Other Pertinent Info: Summary of information relevant to treating heart failure, excluding data already extracted.
Advanced Directive: Including name of proxy, code status, and other goals of care information.

Output Structure: Organize your findings in a JSON object format. Each heart failure element listed below should be a key, with the corresponding value being a list of all of the extracted information.

Example JSON output structure:

```
{
  "Type of Heart Failure": ["HFrEF"],
  "NYHA Class": ["NYHA III-IV"],
  "Ejection Fraction": ["55% from echo on 01/12/2023"],
  "Creatinine": ["Cr 1.29 w/ baseline approx .9-1. Discharge Cr 1.31.", "Cr on discharge was 1.31"],
  "Dry Weight": ["The patient's discharge weight was 187 lbs (84.8 kg) from initial wt of 197 lbs (89.4 kg)"],
  "History of Arrhythmias or VTach": ["The patient has AF w RVR (according to H&P on 06/05/2022)"],
  "Cardiac Medications": [
    "Bumex 1mg daily",
    "Carvedilol 12.5mg BID"
  ],
  "Cardiac Medication Narrative": ["Patient is currently on Bumex, Carvedilol. He was discharged on a regimen of PO Torsemide 20 mg."],
  "Precipitating Events": ["Likely etiology of exacerbation was IVF during recent OSH hospitalization for GIB."]
  "HF Trajectory": ["mild worsening of patient’s congestive heart failure"],
  "Comorbidities": ["amyloid cardiomyopathy, pHTN, HLD, DVT (not on AC), bladder CA s/p TURBT", "AS, AF not on AC (hematuria), DVT"].
```

````
  ...
}
```

If no heart failure content is present in the note, simply respond with "NO HF
CONTENT AVAILABLE".
````

Abstraction Prompt

````
You are a cardiologist tasked with summarizing a patient's heart failure medical
history from their chart upon their admission to the hospital today, {index_date}.
Your sources are multiple electronic health record (EHR) notes, structured as a
dictionary with the format {{"Heart Failure Information Category":
EXTRACTED_INFO_DICT}}. In this dictionary, EXTRACTED_INFO_DICT is a nested
dictionary where keys are descriptions of the source note with note type and date,
and values are the extracted text related to heart failure.

Please provide a comprehensive summary for each of the following Heart Failure
Information Categories by synthesizing information from the various notes.

Type of Heart Failure: Identify whether the heart failure is systolic, diastolic,
etc.
NYHA Class: Class of heart failure indicating severity.
Ejection Fraction: For ejection fraction, mention the value from the last
transthoracic echocardiogram, the most recent value (if different), and the
historical range with dates. (e.g. "most recent EF from last TTE (6/1/2022) is 25%;
last discharge note (6/6/2022) reports EF of 30%
Dry Weight: For dry weight, note the most recent lowest measurement post-discharge,
with the date and note type. Also provide the range of recorded weights. Describe
the context in which the dry weight is recorded (e.g. "dry weight on last discharge
was 185 lbs (180-190 lbs from 1/21-7/22)")
Creatinine: If one of the notes explicitly mentions baseline Creatinine, report it.
Otherwise, report the most recent value along with the date and note type (e.g.,
recent discharge), and separately detail the range of values over time. Mention the
context in which the Cr was taken such as on admission, on discharge, etc. (e.g.
"B/l Cr is 1.7 according to last discharge note (range of 1.7-1.9 from 6/21-7/22)"
)
NT-proBNP / BNP: Report the most recent value along with the date and note type,
and separately detail the range of values over time.
Type of Cardiomyopathy: Describe whether ischemic, non-ischemic, or both.
Cause of Cardiomyopathy: Describe whether due to CAD, autoimmune disorders, etc.
History of Coronary Interventions: Bulleted list of cardiac procedures, such as
coronary angiography, stents, CABG, right heart cath, pacemaker/ICD placement, TAVR
or other valve replacement surgeries, congential heart disease procedures,
cardioversion, heart transplant, cardiac biopsies; Include pertinent results from
recent angiogram if present.
````
````

Recent TTE Findings: Description of most recent and pertinent findings, including EF; Note any issues with obtaining high quality results if present; Include date of TTE.
History of Valvular Disease: Description of history of valvular disease, such as regurgitation or stenosis.
History of Arrhythmias or VTach: Description of history of Afib, Aflutter, Vtach, VT arrest, or Vfib, including pertinent interventions.
Pacemaker or ICD Presence: Details about any ICD, including whether CRT or dual chamber.
Diuretic Medications and Dosage: Bulleted list of diuretics with dosage and frequency that patient is on. Focus on home meds and med changes during admissions.
Cardiac Medications and Dosage: Bulleted list of medications, dosages, and frequency that patient takes. include beta blockers, ace inhibitors, ARB, ARNI, SGLT2 inhibitors, aldosterone antagonists, hydralazine, nitrates, anticoagulants if present. Focus on home meds and med changes during admissions.
Cardiac Medication Narrative and Changes: Summarize the cardiac medication plan, including current meds, side effects, contraindications, and recent changes in cardiac medications with reasons for the changes. Write as a paragraph with shorthand (not a bulleted list) and make sure to include dates of medication changes. Describe reasons for why the patient may/may not be on goal-directed medical therapy, if mentioned. Clarify meds taken in hospital (with date) vs at home. When describing the plan, clearly differentiate between changes for inpatient stay vs changes that will persist post discharge. (e.g. "The patient was started on metoprolol succinate 50 mg daily during recent hospital admission. Plavix was held due to hematoma and restarted post discharge. Recent office visit note on 6/7/2022 reports that cardiologist has tried increasing doses of metolazone 2-3 times a week, but despite a combination of 10 mg of metolazone and 200 mg of torsemide his weight is continuing to go up.")
Outpatient Cardiologist(s): Names of cardiologists seen in an outpatient setting. Do NOT extract the attending cardiologist in the hospital)
Electrophysiologist(s):  Names of electrophysiologists seen in an outpatient setting.
Primary Care Physician (PCP): Patient's outpatient PCP.
Precipitating Events for HF: Summarize known reasons for worsening HF, if present (e.g. "Patient reports not taking at home meds"). Do NOT mention reason for admission if it is unrelated to pt's heart failure.
HF Trajectory: Overall disease trajectory (stable or worsening).
Most Recent HF Exacerbation: Date and description of the most recent hospitalization for HF exacerbation.
Comorbidities: Other pertinent conditions that the patient has. Write as a "one-liner", not as a bulleted list.
Advanced Directive: Including name of proxy, code status, and other goals of care information.
Other Pertinent Info: Summarize other information relevant to diagnosis and management of the patient's heart failure, excluding data already mentioned.

Only include information pertinent to managing heart failure in the future. You do NOT need to include transient information about the patient's prior hospital stays if it is not pertinent to future management.

The output summary should be concise and use medical shorthand / abbreviations.

Prioritize more recent information. Note that the input data includes both (1) the date of each note and (2) dates documented within each note. Base the "most recent" status on the original date of the data, not the note date. For example, the most recent note could refer to an EF from an echo that is outdated. Report the most recent value, not the value reported in the most recent note.

For each piece of information, clarify the context in which it was recorded (e.g. from discharge summary, echo report, admission note, etc.)

Highlight any inconsistencies or contradictions in the extracted data, clearly distinguishing between historical and recent updates, especially if the former may be outdated.

If a category lacks data, state "NA". Each piece of data cited in the summary should be referenced using a numerical order (e.g., [1,2]). At the end, provide a list of cited notes formatted as [Number] Note Type with Chunk Number, Note Date (e.g., [1] Discharge Note #1, 1/13/2021).

**Supplementary Note S3. Survey Design for Chart Review–Based Evaluation.** This note outlines the 19-item questionnaire used to assess the summaries via expert chart review.

**How would you classify this patient's cardiomyopathy?**

* must provide value

- Ischemic
- Non-ischemic
- Combined
- Not enough information available in summary and EHR

reset

**Source you looked at to answer the question?**

* must provide value

- Summary
- EHR
- Not enough information available in summary and EHR

reset

**Does this patient currently have moderate or severe valvular disease (regurgitation or stenosis)?**

- Yes
- No

reset

**Source you looked at to answer the question?**

- Summary
- EHR

reset

**Has this patient had an echocardiogram within the last 6 months (i.e. since March 1 2023)?**

- Yes
- No

reset

**Source you looked at to answer the question?**

- Summary
- EHR

reset

**Does this patient have any of the following known contraindications for ACE inhibitors?**

- **pregnancy**
- **breastfeeding**
- **severe renal failure due to ACE-I**
- **persistent hyperkalemia**
- **persistent hypotension**
- **allergy to ACE-I**
- **patient declined**

**ONLY use the criteria listed above in making this determination.**

- Yes
- No

reset

**Source you looked at to answer the question?**

- Summary
- EHR

reset

**Does this patient have any known history of the following arrhythmia disorders?**

- **Afib**
- **Aflutter**
- **Vtach**
- **Vfib**

**ONLY use the criteria listed above in making this determination.**

◯ Yes

◯ No

reset

**Source you looked at to answer the question?**

◯ Summary

◯ EHR

reset

**Does this patient have any history of the following cardiac procedures?**

- **CABG**
- **coronary angiography**
- **right heart cath**
- **pacemaker/ICD placement**
- **TAVR or other valve replacement surgeries**
- **congenital heart disease procedures**
- **heart transplant**
- **cardioversion**
- **cardiac biopsy**

**ONLY use the procedures listed above in making this determination.**

◯ Yes

◯ No

reset

**Source you looked at to answer the question?**

◯ Summary

◯ EHR

reset

**Which of the following criteria for ICD does the patient meet? Select all that apply.**

☐ EF <= 35% AND NYHA class II/III

☐ presence of NSVT with prior MI

☐ EF <=40% with persistent Vtach or Vfib history

☐ None of the above

**Source you looked at to answer the question?**

◯ Summary

◯ EHR

reset

| | |
|---|---|
| **What is this patient's most recent dry weight?**<br>__________ | **Source you looked at to answer the question?**<br>◯ Summary<br>◯ EHR<br>◯ Not enough information available in summary and EHR<br>reset |

| | |
|---|---|
| **Does this patient follow a cardiologist in the outpatient setting?**<br>◯ Yes<br>◯ No<br>reset | **Source you looked at to answer the question?**<br>◯ Summary<br>◯ EHR<br>reset |

| | |
|---|---|
| **Please select which of the following drug classes of Goal Directed Medical Treatment that the patient is prescribed. Select all that apply:**<br>☐ Beta Blockers<br>☐ Renin-angiotensin system inhibitor (angiotensin converting enzyme [ACE] inhibitor, single-agent angiotensin II receptor blockers [ARB], OR angiotensin receptor-neprilysin inhibitor [ARNI]) (eg. Entresto is an ARNI) (ANY of these options)<br>☐ Sodium-glucose cotransporter 2 (SGLT2) inhibitor (eg. Jardiance/empagliflozin)<br>☐ Mineralocorticoid Receptor Antagonist (eg. spironolactone/aldactone)<br>☐ Hydralazine AND nitrates (must be on both)<br>☐ None of the above | **Source you looked at to answer the question?**<br>◯ Summary<br>◯ EHR<br>◯ Combination of both summary and EHR<br>reset |

| | |
|---|---|
| **Has this patient had any changes in diuretic dose or diuretic medication type within the last 2 months (i.e. since July 1st 2023)?**<br>◯ Yes<br>◯ No<br>reset | **Source you looked at to answer the question?**<br>◯ Summary<br>◯ EHR<br>reset |

| | |
|---|---|
| **The following summary applies to two questions below (Part A and Part B):**<br><br>**The patient presents to the hospital complaining of shortness of breath and is found to be in an acute decompensated heart failure exacerbation. Their HR and blood pressure are within their normal limits.** | |
| **Part A. Assuming they have been taking their typical diuretic dose as prescribed, what medication, dose, and route would you consider giving in the emergency room?**<br><br>Medication<br><br>Dose<br><br>Route | **Source you looked at to answer the question?**<br><br>○ Summary<br>○ EHR<br><br>reset |
| **Part B. They are being admitted into the hospital with stable vitals and no bleeding issues. Which anticoagulation would you prescribe while they are hospitalized?**<br><br>○ Order anticoagulant prescribed at home<br>○ Patient is not on home anticoagulant. Order prophylactic SQH or Lovenox<br><br>reset | **Source you looked at to answer the question?**<br><br>○ Summary<br>○ EHR<br><br>reset |

**Supplementary Note S4.** Instructions provided to physicians

General Instructions

- You will be provided with eight cases and tasked to complete a questionnaire for each case. The questionnaire will include questions about the patient's medical history and how that history will shape your plan for the patient.
- To help you answer the questions, you will have access to either (a) the electronic health record (EHR) for the patient or (b) the EHR and a summary of the patient's heart failure history. The questionnaires need to be completed using the provided sequence of patients.
- Please complete the questionnaire provided below based on this patient's information. You can use all information in a patient's chart up until the time of arrival to the hospital for the specified admission date (i.e., you can even include cardiology and PCP notes on the same index date written before arrival to the hospital, but you should NOT use ER notes or the H&P for the current admission). Please be careful when using the search tool to look up information, as this may surface information from after the patient's date of admission.
- Make sure to open the patient's chart using "patient lookup" instead of opening an encounter.
- Before you begin the questionnaire, we ask that you create filters for the notes, medications, and cardiology tab to filter by the index date. Instructions to do this can be found here.
- You should also NOT use any notes from Care Everywhere. Before you begin the questionnaire, we also ask that you create a filter to remove Care Everywhere notes. Instructions can be found here.
- We are trying to simulate a real chart review for an actual patient. Taking this into consideration, we ask you to complete these survey questions accurately and efficiently, as if you were reviewing your own patient's chart.
- We will time how long it takes you to complete the questionnaire for each patient. Only begin when you are ready. You cannot stop once you start a questionnaire for a single patient. When complete, press the finish button. If you have not completed the questionnaire after thirty minutes, it will automatically end. When you finish, you will NOT immediately be taken to the questionnaire for the next patient. Instead, you'll be taken to an intermediate screen and you can decide to advance to the next set of questions when you are ready.
- All questions in the questionnaires are mandatory.
- After questionnaires where you use the summary, you will be asked to fill out a few questions about the summary. These questions are not timed and are not mandatory.
- Once you are finished with all cases, please fill out a feedback survey describing your experience.

Instructions for evaluation with only EHR

- Please complete the questionnaire based on chart review in Epic for this patient.
- Please make sure to perform all filtering of the EHR before you start filling out the questionnaire.

Instructions for evaluation with EHR and summary

- You will be provided with a summary of this patient's heart failure. You also have access to EPIC.
- Start answering the questionnaire by first referring to the summary to answer the questions. If information is missing from the summary or you are not confident in the information in the summary, you can refer to the EHR to answer the question. You can navigate to relevant notes in the EHR using notes cited in the summary.
- Please familiarize yourself with the layout of the summary before you begin. The summary is split into sections outlining different aspects of the patient's heart failure history. Source notes where the information is pulled from are cited at the bottom of the summary. You can find a summary for an example patient here.
- Please click the button below each question to specify which source you used to answer the question (either Summary or EHR). If at any time you have to revert back to the EHR, select "EHR" as the source of information.
